# Eleven Routine Clinical Features Predict COVID-19 Severity

**DOI:** 10.1101/2020.07.28.20163022

**Authors:** Kai Zhou, Yaoting Sun, Lu Li, Zelin Zang, Jing Wang, Jun Li, Junbo Liang, Fangfei Zhang, Qiushi Zhang, Weigang Ge, Hao Chen, Xindong Sun, Liang Yue, Xiaomai Wu, Bo Shen, Jiaqin Xu, Hongguo Zhu, Shiyong Chen, Hai Yang, Shigao Huang, Minfei Peng, Dongqing Lv, Chao Zhang, Haihong Zhao, Luxiao Hong, Zhehan Zhou, Haixiao Chen, Xuejun Dong, Chunyu Tu, Minghui Li, Yi Zhu, Baofu Chen, Stan Z. Li, Tiannan Guo

**Author notes:** Correspondence (Y.Z.); (B.C.); (Z.L.); (T.G.). co-first (Kai Zhou, Yaoting Sun, Lu Li, Zelin Zang, Jing Wang, Jun Li, Junbo Liang).

## Abstract

Severity prediction of COVID-19 remains one of the major clinical challenges for the ongoing pandemic. Here, we have recruited a 144 COVID-19 patient cohort consisting of training, validation, and internal test sets, longitudinally recorded 124 routine clinical and laboratory parameters, and built a machine learning model to predict the disease progression based on measurements from the first 12 days since the disease onset when no patient became severe. A panel of 11 routine clinical factors, including oxygenation index, basophil counts, aspartate aminotransferase, gender, magnesium, gamma glutamyl transpeptidase, platelet counts, activated partial thromboplastin time, oxygen saturation, body temperature and days after symptom onset, constructed a classifier for COVID-19 severity prediction, achieving accuracy of over 94%. Validation of the model in an independent cohort containing 25 patients achieved accuracy of 80%. The overall sensitivity, specificity, PPV and NPV were 0.70, 0.99, 0.93 and 0.93, respectively. Our model captured predictive dynamics of LDH and CK while their levels were in the normal range. This study presents a practical model for timely severity prediction and surveillance for COVID-19, which is freely available at webserver https://guomics.shinyapps.io/covidAI/.

## INTRODUCTION

The ongoing COVID-19 pandemic, caused by the SARS-CoV-2 virus, poses an unprecedented public health crisis to the entire world. The basic reproductive rate (R_0_) of 2.0-2.5 in COVID-19 is higher than in SARS (1.7–1.9) and MERS (<1) ^1^. To date, SARS-CoV-2 has spread to more than 200 countries and regions around the world, with over 16 million individuals infected. Most confirmed cases are classified as mild or moderate, while the other 14% and 5% are severe and critical cases, respectively ^2^. These patients need to be hospitalized and monitored with intensive care to prevent deterioration of the disease that may lead to fatality without timely diagnosis and treatment.

Currently, diagnosis of COVID-19 mainly depends on the virus RNA test of SARS-COV-2 ^3^. This is a qualitative test showing whether the patient is infected or not ^4^. Besides, computed tomography (CT) is a complementary strategy for COVID-19 diagnosis by showing the image of pneumonia symptoms in lungs ^5^. As a physical image entity, CT can supplement with objective observations to indicate the severity of COVID-19 patients in an AI-assisted model ^6^. This helps stratify the severity of COVID-19 at an early stage of disease progression. However, about 20% of COVID-19 patients show no obvious imaging changes in the lung^7^, bringing difficulties to physicians when making decisions for suitable clinical treatments. To better evaluate the disease conditions of COVID-19 patients, routine laboratory tests including but not limited to complete blood cell counts, blood laboratory metabolomics tests are taken regularly by physicians. Physicians then make clinical decisions and prescribe treatments accordingly. However, this is laborious and sometimes biased, depending on the physician’s own experience, especially when facing with such a heavy medical burden in the pandemic. Therefore, automatic integration and interpretation of routine laboratory indexes in an unbiased way will make a pivotal role in severity stratification and prognosis evaluation for COVID-19 patients.

Since the outbreak of the COVID-19 epidemic, research focusing on building severity prediction models of COVID-19 based on related features have been reported extensively to support medical decision making^8^. Several clinical features, such as age, gender, lactic dehydrogenase (LDH), C-reactive protein (CRP), and lymphocyte count, have been reported to be highly correlated with the severity of COVID-19 patients ^8^. Recently, a Chinese team found that three key features (LDH, CRP and lymphocyte) can be used to predict the mortality of COVID-19 with over 90% accuracy ^9^. Although all models have been reported with great predictability, they have also raised concerns of bias in prediction because of key factors including poor selection of samples, inappropriate statistical methods, *etc* ^*8*^. In a paper published by Liang et al, a risk scoring algorithm based on some key characteristics of COVID-19 patients at the time of admission to the hospital was developed, which may assist in predicting a patient’s risk of developing into critical illness^10^. More recently, they also built a survival model to allow early prediction of critical cases by comorbidity and several laboratory indexes ^11^. These models are mainly built on a dataset either containing static information at certain time points, for example, upon admission or discharge, mostly when the patients were in severe or critical status. However, the time from symptom onset to admission varied in a large range due to different local medical resources and personal medical treatment intention. In contrast, the time of onset of symptoms as a starting point is more objective and reasonable. During the pandemic, personal physical condition and laboratory indicators are constantly changing, however, few models make use of longitudinal measurement to predict disease severity. Therefore, it remains challenging to stratify the risk rank of a patient when she/he is in the transient stage of mild to the early severe stage, due to the lack of robust tests to detect the impalpable and subtle changes inside the patient.

In this study, we built a machine learning model based on the longitudinal measurement of 124 clinical factors in a retrospective cohort containing 144 COVID-19 patients to predict the disease severity. 11 key clinical factors were featured as a panel to be highly associated with COVID-19 severity, instead of isolated features. The model achieved an overall 93% accuracy in distinguishing severe patients among the infected cases in all dataset. The prediction model is freely accessible online as a Shiny Server App (https://guomics.shinyapps.io/covidAI/) to facilitate the early detection of the patients.

## RESULTS AND DISCUSSION

### Clinical characteristics of the study cohort

A total of 841 patients have been screened with the SARS-CoV-2 nucleic acid test from January 17 to March 10, 2020 in Taizhou Hospital, with 144 patients with positive virus RNA (COVID-19) and 697 non-COVID-19 individuals (Figure 1A). From the non-COVID-19 group, outpatients and patients lacking chest CT results were excluded. 65 patients were recruited as the control group, with their epidemiological information collected. Meanwhile, 144 COVID-19 patients were stratified into severe (N = 36) and non-severe (N = 108) patients based on the clinical diagnosis guideline ^7^ (Figure 1A). 124 types of measurements from 17 categories over 52 days were collected and manually curated for these 144 COVID-19 patients, resulting in a data matrix containing of 3,065 readings for 124 types of measurements (3065*124). The 17 categories of data included basic information, clinical symptoms and signs, chest CT results and laboratory tests, as detailed in Table S1 that were regularly recorded during their hospitalization.

**Figure 1.**
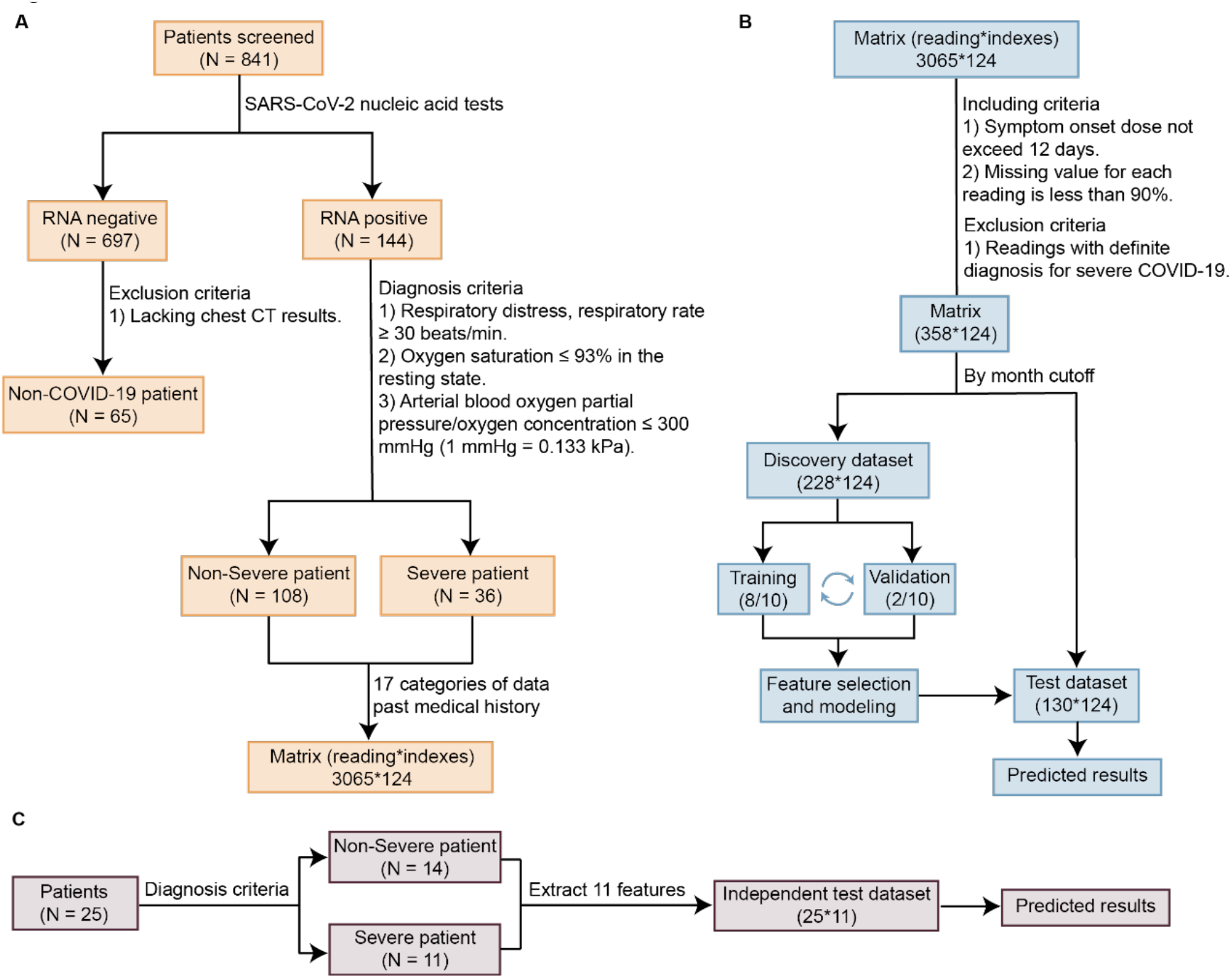
The study design and modeling workflow. **(A)** The COVID-19 patient cohort. **(B)** The workflow of the modeling process. (**C**) Independent test patient cohort.

This cohort has 55.6% male in the severe COVID-19 group and 52.8% male in the non-severe COVID-19 group. The median age was 55.0 years for the severe group, and 44.5 years for the non-severe group. The median BMI was 25.5 in the severe group and 23.8 in the non-severe group (Table 1). These parameters are in consistent with previous observations ^12^.

**Table 1.** Demographic and clinical characteristics of the patients

| Characteristic | non-COVID-19 (N=65) | COVID-19 |  |  | P value |
| --- | --- | --- | --- | --- | --- |
|  |  | Non-severe (N=108) | Severe (N=36) | All (N=144) |  |
| Gender - no. <sup>a</sup> (%) |  |  |  |  |  |
| Male | 41 (63.1) | 57 (52.8) | 20 (55.6) | 77 (53.5) | 0.7723 |
| Female | 24 (36.9) | 51 (47.2) | 16 (44.4) | 67 (46.5) |  |
| Age - yr. <sup>b</sup> |  |  |  |  |  |
| Median (IQR) | 36.0 (21.5-52.0) | 44.5 (37.0-54.0) | 55.0 (47.8-65.0) | 47.0 (38.0-56.0) | 0.0001 |
| Range | (3.0-79.0) | (4.0-86.0) | (33.0-79.0) | (4.0-86.0) |  |
| BMI, kg/m<sup>2</sup> |  |  |  |  |  |
| Median (IQR) | 22.0 (20.5-26.2) | 23.8 (21.5-26.1) | 25.5 (24.1-27.2) | 24.4 (22.0-26.4) | 0.0050 |
| Range | (14.0-32.9) | (16.0-31.2) | (21.6-31.3) | (16.0-31.3) |  |
| Onset to severe, days |  |  |  |  |  |
| Median (IQR) | NA | NA | 9 (5-12) | NA | NA |
| Range | NA | NA | (2-26) | NA |  |
| Admission to severe, days |  |  |  |  |  |
| Median (IQR) | NA | NA | 2 (1-3) | NA | NA |
| Range | NA | NA | (0-7) | NA |  |
| Length of stay, days |  |  |  |  |  |
| Median (IQR) | 4 (3-6) | 20 (13-27) | 23 (19-32) | 22 (13-28) | 0.0154 |
| Range | (1-31) | (6-44) | (9-40) | (6-44) |  |
| Smoking - no. (%) | 5 (7.7) | 11 (10.2) | 2 (5.6) | 13 (9.0) | 0.5182 |
| Familial aggregation - no. (%) | 7 (10.8) | 43 (39.8) | 11 (30.6) | 54 (37.5) | 0.3203 |
| Symptoms and signs - no. (%) |  |  |  |  |  |
| Fever | 35 (53.9) | 70 (64.8) | 34 (94.4) | 114 (79.2) | 0.0006 |
| Pharyngalgia | 14 (21.5) | 15 (13.9) | 2 (5.7) | 17 (11.8) | 0.2405 |
| Cough | 38 (58.5) | 47 (43.5) | 18 (50.0) | 65 (45.1) | 0.4985 |
| Expectoration | 25 (38.5) | 19 (17.6) | 7 (19.4) | 26 (18.1) | 0.8025 |
| Muscle soreness | 1 (1.5) | 4 (3.7) | 2 (5.6) | 6 (4.2) | 0.6399 |
| Headache | 1 (1.5) | 9 (8.3) | 7 (19.4) | 16 (11.1) | 0.1208 |
| Diarrhea | 24 (36.9) | 3 (2.8) | 3 (8.3) | 6 (4.2) | 0.1652 |
| Chest tightness | 4 (6.2) | 7 (6.5) | 4 (11.1) | 11 (7.6) | 0.4676 |
| Heart rate |  |  |  |  |  |
| Median (IQR) | 96 (86-107) | 82 (75-90) | 84 (75-95) | 82 (75-91) | 0.5106 |
| Range | (72-126) | (57-147) | (57-106) | (57-147) |  |
| Respiratory rate |  |  |  |  |  |
| Median (IQR) | 19 (19-20) | 19 (18-20) | 19 (18-20) | 19 (18-20) | 0.6265 |
| Range | (18-28) | (12-26) | (12-22) | (12-26) |  |
| Comorbidities - no. (%) |  |  |  |  |  |
| Hypertension | 7 (10.8) | 14 (13.0) | 8 (22.2) | 22 (15.3) | 0.1811 |
| Diabetes | 6 (9.2) | 9 (8.3) | 5 (13.9) | 14 (9.7) | 0.3397 |
| Cardiovascular disease | 2 (3.1) | 1 (0.9) | 2 (5.6) | 3 (2.1) | 0.1543 |
| Cerebrovascular disease | 4 (6.2) | 2 (1.9) | 1 (2.8) | 3 (2.1) | 1.000 |
| Chronic bronchitis | 3 (4.6) | 4 (3.7) | 0 (0.0) | 4 (2.8) | 0.5721 |
| Tuberculosis | 0 (0.0) | 4 (3.7) | 0 (0.0) | 4 (2.8) | 0.5721 |
| Malignant tumor | 2 (3.1) | 1 (0.9) | 1 (2.8) | 2 (1.4) | 0.4388 |
| Thyroid disease | 2 (3.1) | 2 (1.9) | 3 (8.3) | 5 (3.5) | 0.0998 |
| Hepatitis | 1 (1.5) | 5 (4.6) | 2 (5.6) | 7 (4.9) | 1.0000 |
| Chronic kidney disease | 1 (1.5) | 1 (0.9) | 1 (2.8) | 2 (1.4) | 0.4388 |
| Digestive system disease | 3 (4.6) | 6 (5.6) | 1 (2.8) | 7 (4.9) | 0.6803 |
| **Chest CT <sup>c</sup> - no. (%)** |  |  |  |  |  |
| Involvement of chest radiographs | 47 (72.3) | 106 (98.1) | 36 (100.0) | 142 (98.6) | 1.0000 |
| Unilateral lung | 14 (21.5) | 28 (25.9) | 3 (8.3) | 31 (21.5) | 0.0261 |
| Bilateral lung | 33 (50.8) | 65 (60.2) | 31 (86.1) | 96 (66.7) | 0.0043 |
| Pulmonary plaques | 13 (20.0) | 52 (48.2) | 24 (66.7) | 76 (52.8) | 0.0539 |
| Ground-glass opacity (GGO) | 2 (3.1) | 34 (31.5) | 15 (41.7) | 49 (34.0) | 0.2640 |
| Pulmonary fibrosis | 3 (4.6) | 7 (6.5) | 1 (2.8) | 8 (5.6) | 0.6796 |
| Pulmonary consolidation | 11 (16.9) | 23 (21.3) | 9 (25.0) | 32 (22.2) | 0.6434 |

<sup>a</sup> no. (%): number.
<sup>b</sup> yr.: year.

The most common symptom at disease onset was fever (64.8% in non-severe and 94.4% in the severe group), followed by cough and pharyngalgia (Table 1). Before admission, 40.2% of patients had underlying diseases and the ratio is substantially higher in the severe group (50.0%) than that in the non-severe group (37.0%). Hypertension (15.3%) and diabetes (9.7%) were common comorbidities.

Upon admission, 142 (98.6 %) COVID-19 patients had image changes in chest CT. Pulmonary plaque was the top 1 abnormal pattern (52.8%), with ratios of abnormality of 66.7% for severe patients and 48.2% for non-severe patients, respectively. The second abnormal pattern was ground-glass like symptoms (severe vs. non-severe was 41.7% vs. 31.5 %). Ratios of pulmonary fibrosis and consolidation did not show a statistical difference between severe patients and non-severe ones.

In severe patients, intervals from onset and admission to the diagnosis of severe cases were 9 (5-12) days and 2 (1-3) days. All 144 patients were followed up during their entire course in hospital. In this study, only one patient was admitted to the intensive care unit (ICU) and underwent invasive mechanical ventilation. They were all cured and discharged eventually. The mean length of stay for the patients was 20 days for non-severe group and 23 days for severe group, respectively. In summary, this is a well-annotated and curated COVID-19 cohort with comprehensively and systematically recorded information from the disease onset till convalesce and discharge, which provided the potential for subsequent model construction.

### Feature selection and machine learning

To establish a model for severity prediction, we firstly filtered the matrix including 3065*124 readings for all patients over 52 days in total (Figure 1B). For the first step, the readings with definite diagnosis for severe COVID-19 were excluded. Then the readings recorded from symptom onset to the 12^th^ day for all patients were included for severity prediction by machine learning, in which the ratio of severe verse non-severe was close to 20% prevalence of severe cases according to previous studies ^2^. It is worthy of noting that at this time point (12^th^ day), no patients had been clinically diagnosed as severe COVID-19 cases. After removing readings with more than 90% missing values, a much smaller data matrix containing 358*124 readings remained. The discovery dataset contained 228*124 readings collected from patients admitted before 1 February, 2020, while the test dataset containing 130*124 readings from patients admitted after 1 February, 2020. were generated. Based on the discovery dataset, a machine learning model was built up by cross training and validation (Figure 1B). To further test our model, we also collected an independent dataset from another clinical center, employing the same criteria with the previous one (Figure 1C).

The process of modeling contained three parts, including feature selection, model training and prediction (Figure 2A). In detail, firstly, the missing value for each test item in the discovery dataset was filled with a relevant median value for each gender (F or M). We randomly generated feature combinations and loaded the data with selected features. Then feature selection was performed by using genetic algorithm (GA) ^13^, one of the most advanced and widely used algorithm for feature selection, assisted with 10-fold cross-validation in the discovery dataset including both training (9/10) and validation (1/10) sets. The method of GA which selected a panel of features could avoid being trapped into local optimal solution. As a result, a panel of eleven key clinical factors that are associated with COVID-19 severity was evolved from 124 characteristics, and were put into the ‘Active Feature Pool’.

**Figure 2.**
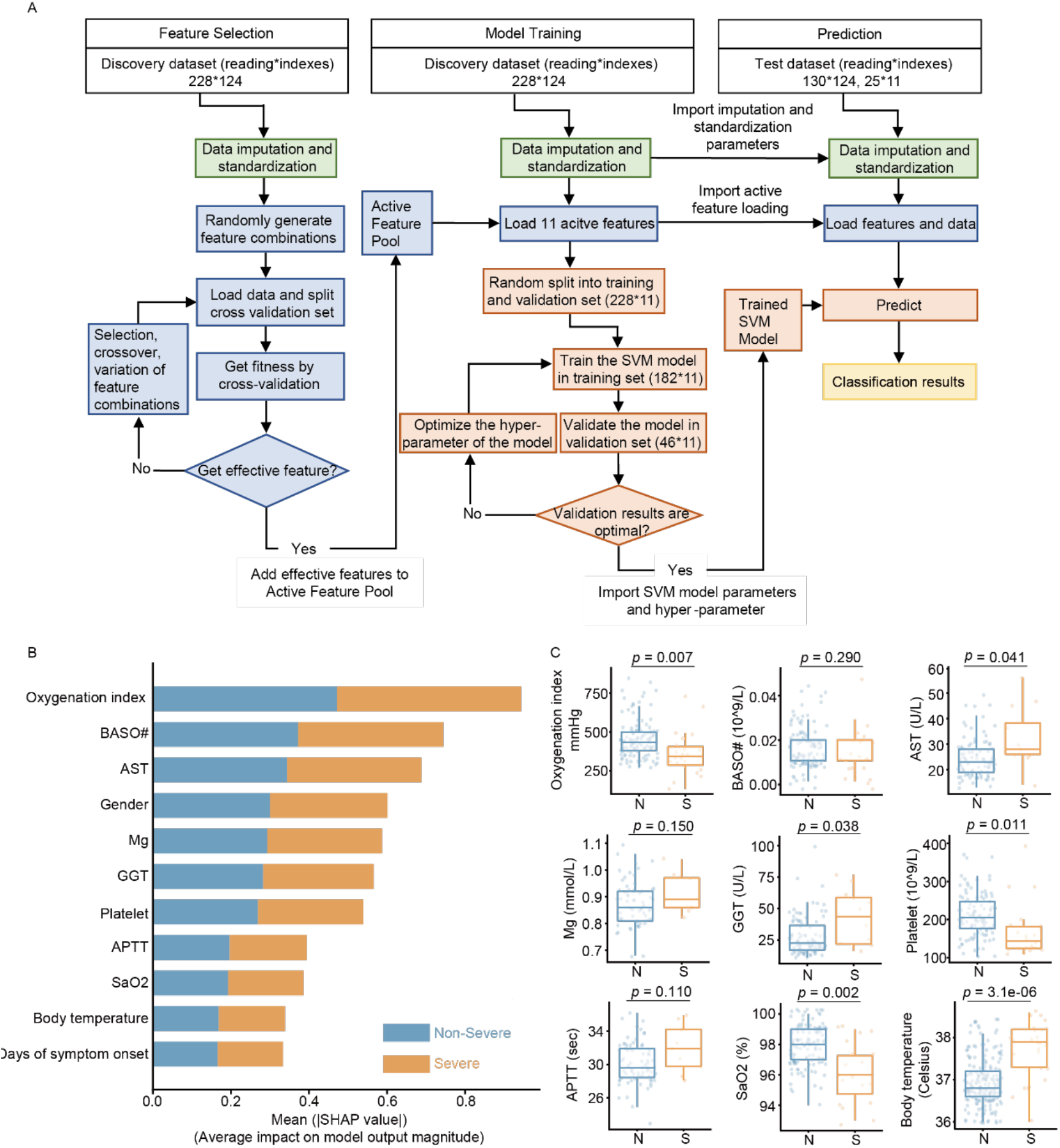
The machine learning model. **(A)** The detailed workflow contains four major steps: i) data preprocessing (green); ii) feature selection (blue); iii) building up the machine learning model (pink); iv) prediction results (yellow). **(B)** The 11 selected key clinical features displayed in two groups according the severity of the disease (blue: non-severe; orange: severe) from the discovery cohort. **(C)** The comparison of each key feature between non-severe and severe COVID-19 patients in the discovery dataset. *P-*value was calculated by the unpaired two-tailed Student’s t-test. N, non-severe; S, severe.

Then we applied the selected features and established the classifier by using the support vector machine (SVM) ^14^. In this step, we set up 10 random seeds and the SVM model was trained in the training set (8/10) and then validated in the validation set (2/10). The hyper-parameters of model and threshold were optimized to maximize accuracy. The threshold was set at 0.45, which was determined by maximizing the correct rate of diagnosis in the validation set. Via this way, a trained SVM model including the eleven active features and classifier was generated for predicting severe cases in the test dataset (130*124) and an independent test dataset (25*11) using the same threshold of 0.45 (Figure 2A).

To interpret the importance of each individual feature applied in the model, SHapley Additive exPlanations (SHAP) algorithm ^15,16^ was performed. The eleven features, oxygenation index, basophil counts (BASO#), aspartate aminotransferase (AST), gender, magnesium (Mg), gamma glutamyl transpeptidase (GGT), platelet counts, activated partial thromboplastin time (APTT), oxygen saturation (SaO2), body temperature and days of symptom onset, were then arranged according to their importance to the model evaluating by SHAP values (Figure 2B). The results showed that oxygenation index was the most important one with a SHAP value of 0.94 in our model. In clinical practice, oxygenation index is also a critical factor to evaluate the state of disease severity.

Furthermore, nine features with continuous records (12 days) were selected and compared in severe and non-severe groups. Their median values (discovery dataset) were calculated and showed in boxplots (Figure 2C). Six of nine features were significantly (p-value less than 0.05) dysregulated. Among them, oxygen index and SaO2 were decreased in severe cases. This was directly associated with pulmonary function. Two selected features, AST and GGT, were statistically up-regulated, indicating that COVID-19 may induce slight hepatic injury in the early stage. Although within the normal range, the count of platelets was reduced significantly in severe cases. Three characteristics, namely APTT, Mg and BASO#, showed no significant change between severe and non-severe COVID-19 patients, a comprehensive comparison was further performed. As shown in Figure S1, readings from four groups of individuals, severe COVID-19, non-severe COVID-19, non-COVID-19 patients with flu-like symptoms and healthy people with physical examination were included for systematic comparison. The data showed that APTT was substantially up-regulated in severe COVID-19 group in the entire dataset including the discovery and the test datasets, and the count of basophils was decreased in all COVID-19 patients compared with healthy group. However, Mg showed no difference across all four groups. To validate the importance of Mg in the model, we used the rest ten features except of Mg and investigated the performance of the model in ROC (Figure S2). The AUC values were the same in training and validation dataset and only decreased 0.01 in test dataset without the feature of Mg, which indicated little contribution of Mg to the model.

The remaining two indicators not shown in the box diagram were days of symptom onset and gender. There is no surprise that the earlier outstood because when the patients were evolving from non-severe to severe status, the symptoms grew worse. Gender was selected as another important feature. In our dataset, male patients were more likely to be infected SARS-CoV-2 than female patients in both non-severe and severe groups, in consistent with the literature reporting that male COVID-19 patients had a worse outcome and that 70% of patients died of COVID-19 were male in an Italy cohort ^17^. The vulnerability of males has also been found in SARS-CoV epidemic in 2003 ^18^. It has been recently found that the plasma concentration of ACE2, a functional receptor for SARC-CoV-2 infection, is higher in men than in women as detected in two independent cohorts ^19^. This may explain the association between gender and fatality rate of COVID-19. Meanwhile, females develop enhanced innate and adaptive immune responses than males did, thus they are less susceptible to kinds of infections of bacterial, viral, parasitic, and fungal origin and malignancies ^20^. In fact, the clinical manifestations of infectious or autoimmune diseases and malignant tumors differ between men and women.

### Severity prediction by machine learning

The model assigned a score (from 0 to 1) to indicate the likelihood of disease severity, and higher score indicated greater severity. The model is described in details in Methods, while the data are shown in the scatter plot (Figure 3A). In this study, samples with a score greater than 0.45 was identified as a severe state. The receiver operating characteristics (ROC) plot achieved an area under the curve (AUC) value of 1.00 and 0.98 for training and validation datasets, respectively (Figure 3B). 224 out of 228 samples were correctly identified with an accuracy of 0.98 for the discovery dataset. The sensitivity, specificity, positive predictive value (PPV) and negative predictive value (NPV) were 0.90, 0.99, 0.96 and 0.99, respectively (Figure 3C).

**Figure 3.**
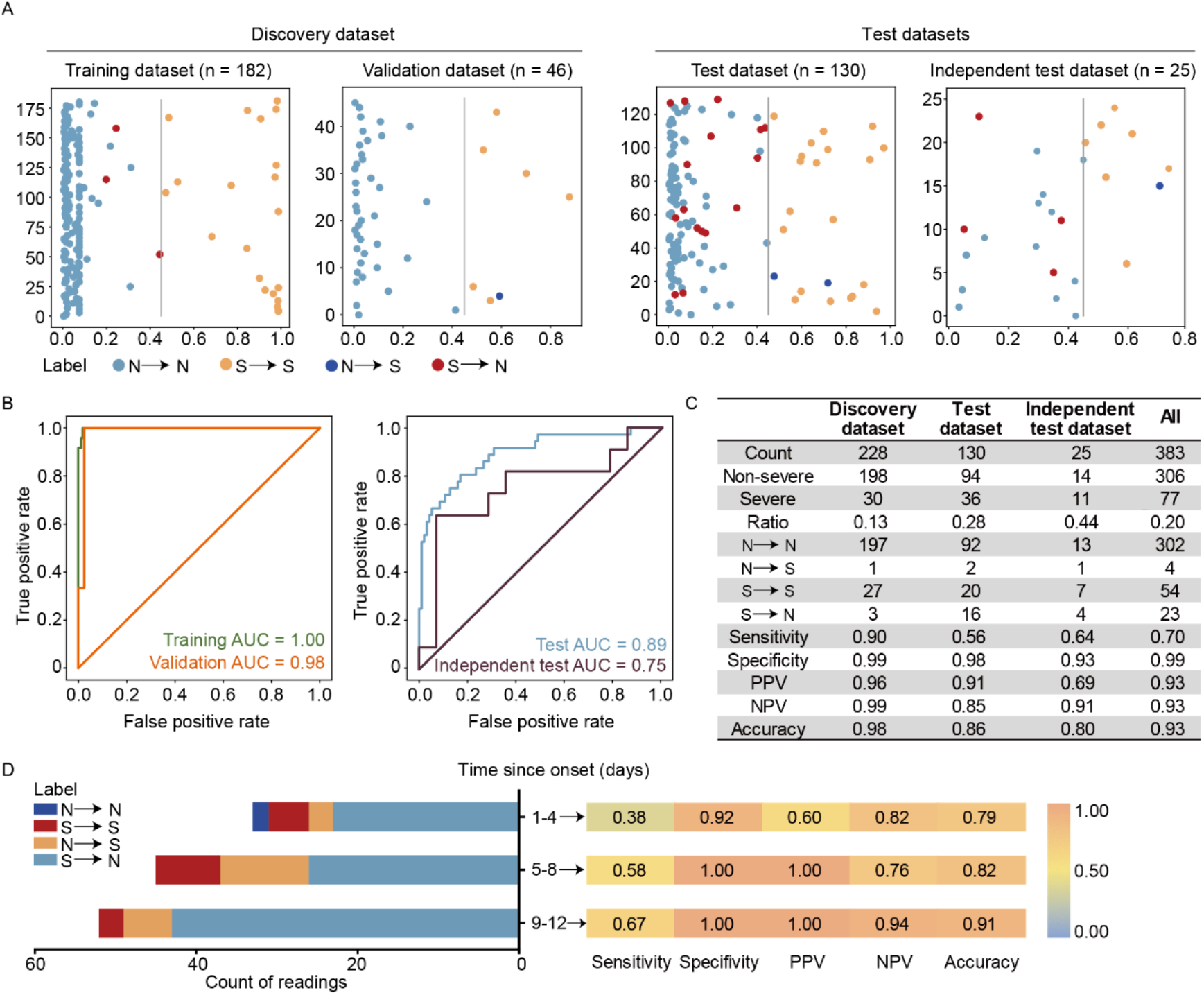
The performance of the predictive model. **(A)** Severe and Non-severe cases are shown as scatter plots in different colors (red: severe; blue: non-severe). X-axis indicates the predicted scores, representing the probability of disease severity for each timepoint. The cutoff of the predicted score was 0.45. Y-axis denotes the indexes of samples. N→S indicates a non-severe case was predicted as a severe case. **(B)** ROC plots of the performance of support vector machine (SVM) for severity prediction. **(C)** Summary of the performance metrics. **(D)** The test dataset was further divided into three longitudinal parts at a four-day interval. The X-axis represents the count of the readings for four types of predicted outcomes in each part (N→N, S→S, N→S, and S→N).

To further evaluate the performance of the 11-clinical feature classifier, we analyzed 130*124 readings in the test dataset and 25*11 readings in the independent test dataset from a different hospital. The classifier achieved AUC value of 0.89 (Figure 3B), and correctly classified 112 of 130 readings with accuracy, sensitivity, specificity, PPV, and NPV of 0.86, 0.56, 0.98, 0.91 and 0.85. Sixteen of 18 incorrectly classified readings were from severe COVID-19 group, especially in early stage of COVID-19 which showed similar clinical signs with non-severe group. Most of the incorrect predictions (10 of 18 readings) were from four COVID-19 patients, indicating individual effect on the model. As to the independent test dataset of much smaller size, the classifier achieved an AUC value of 0.75, and accuracy, sensitivity, specificity, PPV, and NPV of 0.80, 0.64, 0.93, 0.69 and 0.91, respectively (Figure 3C). All incorrectly identified cases in this dataset belonged to early stage of COVID-19 with days of symptom onset less than 8 days.

We also examined the performance of the model from the longitudinal perspective. The test dataset was divided into three parts according to the length of symptom lasting period since disease onset. In general, the longer the disease progressed, the better the model predicted. As the onset time increased, the accuracy of prediction elevated from 0.79 (1-4 days), 0.82 (5-8 days) to 0.91 (9-12 days) in the test set (Figure 3D). There were two incorrectly identified readings in non-severe patients. They were both from the initial three days since onset, probably due to the low oxygen index (less than 380) which was out of the normal range (400-500). As to the part of 9-12 days, three readings of severe group were classified into non-severe group, while two of them were scored with 0.42 and 0.44, which were very close to the threshold.

### Dynamic changes of key clinical features over 7 weeks

The unique curated longitudinal data permitted detailed temporal investigation of specific parameters. We investigated the temporal changes of the eight out of eleven key clinical features in both severe and non-severe COVID-19 patients during the first 40 days since disease onset. In addition, we also analyzed 18 more clinical features (Figure 4) that found to be related to the progression of COVID-19 disease ^8,21,22^.

**Figure 4.**
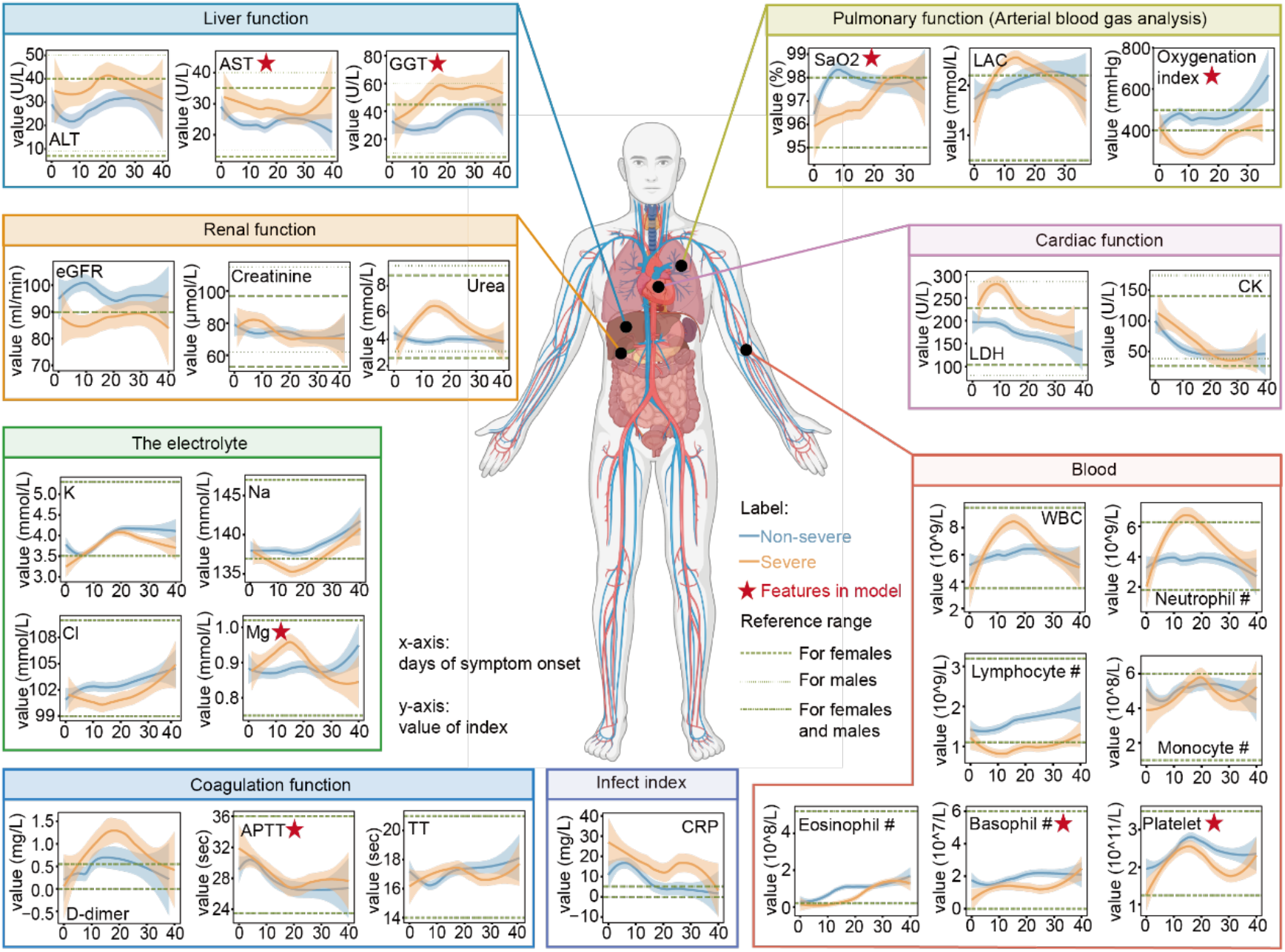
Temporal dynamics of selected clinical indexes in COVID-19 patients. The x-axis represents the time of disease course since symptom onset, and y-axis shows the measured values for each feature over time in severe (orange line) and non-severe (blue line) patients. The solid line was fitted by locally estimated scatterplot smoothing (LOESS). Abbreviations: ALT, alanine transaminase; LAC, lactic acid; eGFR, estimated glomerular filtration rate; LDH, lactate dehydrogenase; CK, creatine kinase; K, potassium; Na, sodium; Cl, chlorine; WBC, white blood cell; TT, thrombin time; CRP, C-reactive protein.

Based on longitudinal changes of blood cell counts (such as platelet), our data showed that the severity of the disease intensified in the second week since symptom onset (Figure 4). The counts of platelets, lymphocytes, eosinophils, and basophils were reduced in the severe group, while neutrophils and WBC were dramatically elevated (Figure 4).

Decreased number of lymphocytes has been reported in association with the COVID-19 severity ^23^, probably due to apoptosis and necrosis of lymphocytes ^24^. Dysregulation of neutrophils was another predominant alteration in the complete blood count test. Neutrophils are recruited early to sites of infection where they kill pathogens (bacteria, fungi, and viruses) by oxidative burst and phagocytosis ^25^.

A significant rise in neutrophil counts features the severity of COVID-19. Increase of neutrophils may induce formation of neutrophil extracellular traps (NETs) which could trigger a cascade of inflammatory reactions ^26^. The latter destroys surrounding tissues, facilitates micro thrombosis, and results in permanent organ injuries to the pulmonary, cardiovascular, and renal systems ^25^, which are three commonly affected organ systems in severe COVID-19^27^.

Eosinophils were drastically reduced after SARS-CoV-2 infection and were merely detectable in the acute stage (Figure 4). The increase of eosinophil in non-severe patients happened earlier than that in severe cases, suggesting that the increase of eosinophils can be the signs for COVID-19 recovery. The predictive value of Eosinophil count observed in our study is supported by a recent independent study ^28^. The count of Basophils is another predictive feature from our model. Basophils are the least abundant granulocytes, representing less than 1% of peripheral blood leukocytes. Recent studies have shown that the count of basophils was reduced in COVID-19 patients ^29^, agreeing with our data. Moreover, an independent study which proposed a multivariate Cox regression model exploring risk factors for lethal COVID-19 patients also nominated progressive increase of basophils ^30^.

Coagulation dysfunction has been found in multiple epidemiologic studies of COVID-19 ^27,31^. Our data showed that platelet counts in blood decreased in severe patients significantly (Figure 4), in consistent with previous meta-analysis reporting low platelet count as a risk factor for COVID-19 severity and mortality ^32^. Furthermore, coagulation related parameters such as D-dimer elevated too, especially in death cases ^31^. Thrombin time (TT) and APTT are often used to monitor the coagulation function of patients, which are mostly increased in the presence of heparin or heparin-like substances.

CRP is an acute-phase protein, expression level of which in blood is associated with the severity of inflammation ^33^. As an indicator of infection, in our dataset, CRP was significantly increased in both mild and severe cases, far beyond the normal range. CRP decreased as the disease progressed and recovered.

We also evaluated functions of organs since it has been reported that SARS-CoV-2 infection systematically induced multi-organ dysfunctions, such as lung, liver, kidney and heart, particularly in severe and critical cases ^34,35^. The commonly used indicators in clinic include pulmonary function indicators including SaO2, blood-gas lactate (LAC) and oxygen index, liver function indicators (GGT, AST and ALT), renal function indicators (eGFR and creatinine) and cardiac function indicator (LDH), were monitored. Arterial blood gas (ABG) analysis is a key approach to evaluate the function of lungs by measuring acidity and the level of oxygen and carbon dioxide, which provides important indexes and direct evidence for indicating pulmonary function and the severity of COVID-19. Our data showed that oxygen-related indexes, namely SaO2 and oxygen index, decreased in severe COVID-19 patients (Figure 4), suggesting a high degree of sensitivity for severity prediction and thus selected as pivotal features in our model. The level of blood-gas LAC reached peak in the first 10 days in severe group, agreeing with the literature ^21^.

Regarding to liver functions, the dynamic change of GGT, AST and ALT showed that hepatic functions got worse around the third week (Figure 4). Moreover, levels of the three indexes were higher in severe patients than in non-severe patients from the beginning of the disease. This may be partly due to the side effect of multi-drug administration according to ALT and GGT level. AST was a predominant feature in our predictive model. which was elevated in the severe group. A multicenter retrospective study including 5771 adult patients reported that AST increased first, followed by ALT in severe cases with liver injury. This alternation was associated with dysregulation of lymphocyte and neutrophil counts ^36^, the latter were selected by our model as well.

Estimated glomerular filtration rate (eGFR) is a direct indicator of renal function and its level in severe patients was significantly lower than that in non-severe patients. On the opposite, the level of urea was elevated in severe group in the early stage (first 10 days), while remained stable in non-severe group. Electrolytes were also closely related to renal function, and most of them were within the reference interval except for sodium and magnesium in severe group (Figure 4C).

LDH is widely expressed in various organs and it has been reported as a key cardiac marker closely associated with COVID-19 severity ^37^. In our study, LDH peaked around the 10th day in severe COVID-19 patients comparing with non-severe cases, although the peak value did not exceed the upper limit of the normal range of LDH for males. Another cardiac indicator, CK, was also in normal range, however, its dynamics differed between severe and non-severe patients.

## CONCLUSION

In summary, we built a customized machine learning model for COVID-19 severity prediction based on the longitudinal measurement of the feature clinical factors over time. The model provided a classifier composed of 11 routine clinical features which are widely available during COVID-19 management. which could predict the prognosis and may guide the medical care of COVID-19 patients. Although some interesting alterations have been found, the major limitation of the present study is the limited patient cohort recruited and the relatively smaller size of it. Therefore, the model still needs to be validated in sizeable cohorts from multiple clinical centers.

## Data Availability

This study presents a practical model for timely severity prediction and surveillance for COVID-19, which is freely available at webserver https://guomics.shinyapps.io/covidAI/.

## ACKNOWLEDGMENTS

This work is supported by grants from Tencent Foundation (2020), National Natural Science Foundation of China (81972492, 21904107, 81672086), Zhejiang Provincial Natural Science Foundation for Distinguished Young Scholars (LR19C050001), Hangzhou Agriculture and Society Advancement Program (20190101A04). This work was also financially supported by National Natural Science Foundation of China (Grant NO.81672086). We thank the patients who enrolled in this study, and the physicians, nurses, and secretaries of the Taizhou Hospital for their important contributions.

## AUTHOR CONTRIBUTIONS

T.G., B.C., J.Liang., S.L., B.S., J.Li., H.C. and Y.Z. supervised the project. T.G. and Y.S. designed the study. K.Z., J.W., M.P., X.Z., Z.Zhou., H.Zhao., J.X., H.Zhu., J.Li., S.C., H.Y., D.L., C.Z., L.H., and X.W. collected clinical data and provided clinical supervision. S.H. organized the data, Y.S., L.L., J.W., K.Z., F.F., X.S., Q.Z., W.G., L.Y., X.S. and H.C. conducted data analysis and visualization. Z.Zang. established machine learning model. X.D., C.T., and M.L. collected the validate data. Data were interpreted and presented by all co-authors. Y.S., L.L., Y.Z., and T.G. wrote the manuscript with inputs from all co-authors.

## MATERIALS AND METHODS

### Patients

From January 17 to March 10 in 2020, we collected clinical and laboratory data for 144 patients in Taizhou Public Hygiene Taizhou Hospital of Zhejiang Province who were diagnosed as COVID-19 infected patients by reverse transcriptase-polymerase chain reaction (RT-PCR) and chest CT according to the Pneumonia Diagnosis and Treatment Scheme for New Coronary Virus Infections (Trial Edition 5, Revision). Clinical data for these patients were curated from the hospital information system (HIS) and included epidemiology, clinical data such as gender, age, BMI, underlying diseases, chest CT, presenting symptoms and length of stay (LOS). Laboratory data included complete blood count, blood chemistry panel, blood clotting test, cytokine panel blood test and arterial blood gas test. Twenty-five independent test readings were collected from Shaoxing People’s Hospital according to the same criteria with the Taizhou test set. This study was approved by the Medical Ethics Committee of Taizhou Hospital and Westlake University, Zhejiang province of China, and informed consent was obtained from each enrolled subject. Besides, the case of minors enrolled in the study was approved by parents and/or legal guardian.

### Laboratory measurements

Blood samples were collected at each time point since admission. 2 mL EDTA-K2 anticoagulant peripheral blood samples were measured for blood routine test using a Sysmex 2100D routine hematology analyzer (Kobe, Japan). Erythrocyte sedimentation rate (ESR) was calculated using the Alifax Test 1 automatic ESR analyzer (UDINE, Italy). Cytokines and lymphocyte subsets were measured using BD FACSCantoTM II within 6 hours. Sodium citrate plasma samples were centrifuged at 3000 rpm for 15 min. Coagulation parameters were determined using a Sysmex CS 5100i automatic hemagglutination analyzer (Kobe, Japan). Arterial blood gas analysis was performed using GEM Premier 3500 (Instrumentation Laboratory, US).

Serum samples were centrifuged at 2500-3000 rpm for 5-10 min for measuring biochemical data including electrolyte, liver and kidney functional proteins, immunoglobulin serial index, blood lipid and glucose, myocardial enzymes(except myoglobin, creatine kinase MB and troponin-I), and infection index (except procalcitonin) using Beckman automatic biochemical analyzer (AU5821). Myoglobin, creatine kinase MB and troponin-I were detected by Beckman automatic immunological analyzer (UniCel DxI-800). Procalcitonin (PCT) levels were determined using the Roche Cobas e411 electrochemiluminescence analyzer (Basel, Switzerland).

Nasopharyngeal and sputum specimens collected during hospitalization were sent to the PCR laboratory (BIOSafety Laboratory II) in a biosafety transportation box. Total nucleic acid extraction from the samples was performed using Nucleic Acid Extraction Kit (Shanghai Zhijiang) and qRT-PCR was performed using a commercial kit specific for 2019-nCoV detection (triple fluorescence PCR, Shanghai Zhijiang, China, NO. P20200105) approved by the China Food and Drug Administration (CFDA).

### Chest radiography

All patients had a chest CT examination at the time of admission. For each patient, the chest CT was performed once a week during the period of her/his hospitalization. A CT examination was also operated at the time of her/his discharge. The points of CT score is defined by the following rules. Based on the lesion involvement and lesion properties, each infected lobe adds one point. Presence of ground-glass opacity adds one point. Two points were added in the presence of consolidation lesions, while three points were added in case of fibrosis lesions. The score was reduced by 0.5 point if the CT is improved compared to the previous CT scan. Otherwise, the score was increased by 0.5 point.

### Machine learning

The data matrix was divided into a discovery dataset and a test dataset according to the admission date. This method defined the samples collected before February 2 as the discovery dataset and the samples collected after February 2 as the test dataset. Feature selection and model training were performed in the discovery dataset. The model was then tested in the test dataset and independent test dataset for evaluation.

The data analysis included four steps: data preprocessing, feature selection, model training and testing. In the data preprocessing step, all missing values were first filled with a median value of all patients. Then as normalization, the mean value of all features in the discovery set was converted to 0 and the standard deviation was converted to 1. The same normalization parameters were applied to the test set. In the feature selection step, the standard GA method in the Python deap library is used. Set the gene locus in the method as the index of the feature, and set the length of the gene chain to 20 (this indicates that the method can select up to 20 different features). Set the crossover probability to 0.3, the mutation probability to 0.5, the number of genes in the population to 500, and the number of iterations to 30. The accuracy rate of the 10-fold cross-validation on the verification set was taken as the fitness of the gene chain. Finally, we selected 11 non-repeated and effective features. In the model training step, the discovery dataset was divided into training set and the validation set. The training set was used to train the model, while the validation set was used to optimize the model parameters.

Afterwards, the test dataset was used to test the model for the prediction accuracy. The SVM model with ‘rbf’ kernal in Python’s sklearn library was employed. The regularization parameter C was set to 1.2. The kernel coefficient gamma was set to ‘auto’. The random state was set to 1. We have optimized the above parameters.

The detection result of patient *i* on a certain feature is *D*_*i,f*_, then the data composed of the selected features of the patient is *D*_*i*_. Then we have 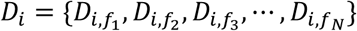 as the number of selected features. Enter *D*_*i*_ into the trained model *M*(⋅), and the model will output a predicted score *S*_*i*_.

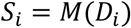

We made the diagnosis based on predicted score *S*_*i*_.

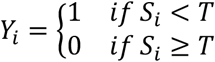

When *Y*_*i*_ is 1, patient *i* is diagnosed as severe. When *Y*_*i*_ is 0, patient *i* is diagnosed as mild. *T* is the threshold for diagnosis.

Due to the heterogeneity of the positive and negative samples, it is not possible to directly use *T* = 0.5 for diagnosis. The threshold *T* is therefore determined by maximizing the correct rate of diagnosis in the validation set, and the same *T* was applied to the evaluation test set.

### Statistical analysis

For statistics analysis, we used R software (version 3.6.3). The comparison of continuous variables between two groups was made using Student’s t-test for normally distributed variables and the Mann-Whitney U statistics for non-normally distributed variables. *P-*value in boxplot was calculated by the unpaired two-tailed Student’s t-test and in violin plot p-value was adjusted by Benjamini & Hochberg. The smooth plot was fitted by locally estimated scatterplot smoothing (LOESS) using the geom smooth function in ggplot. PPV and NPV were adjusted by ratio of severe cases followed by published formula ^38^. SPSS (version 19.0) was used for baseline characteristics.

## SUPPLEMENTARY FIGURE LEGEND

**Figure S1.**
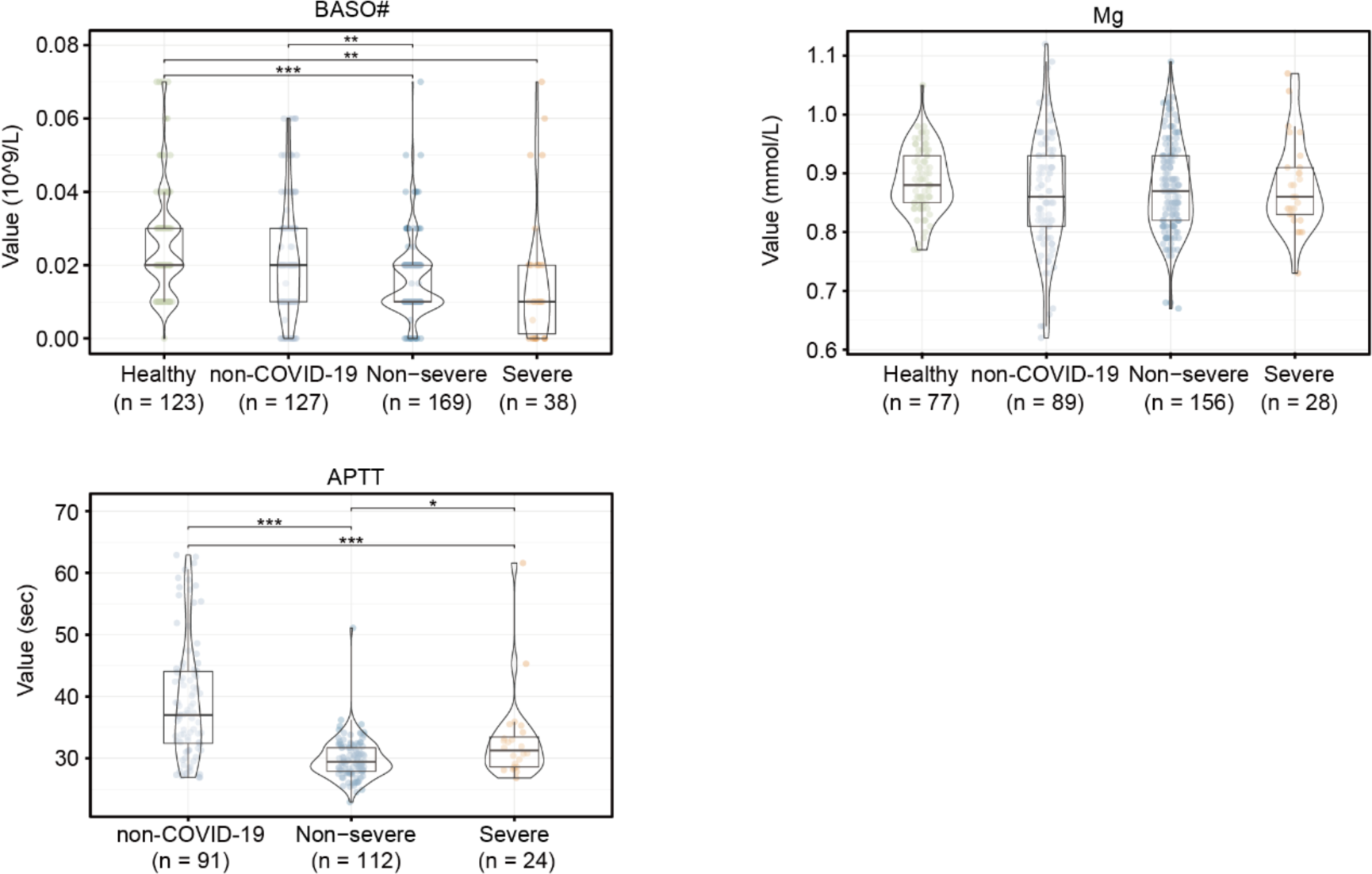
The abundance of selected clinical features in four groups. 146 healthy people with 150 physical examination readings and 65 non-COVID-19 patients with 212 hospitalized detection readings, 108 non-severe COVID-19 patients with 292 readings, and 36 severe COVID-19 patients with 66 readings were collected. We deleted the outliers whose absolute value of Z-score was higher than 3. N refers to the count of data involved. P-value was performed by Yuen’s trimmed means test and was adjusted by Benjamini & Hochberg. p-value: *, <0.05; **, <0.01; ***, <0.001.

**Figure S2.**
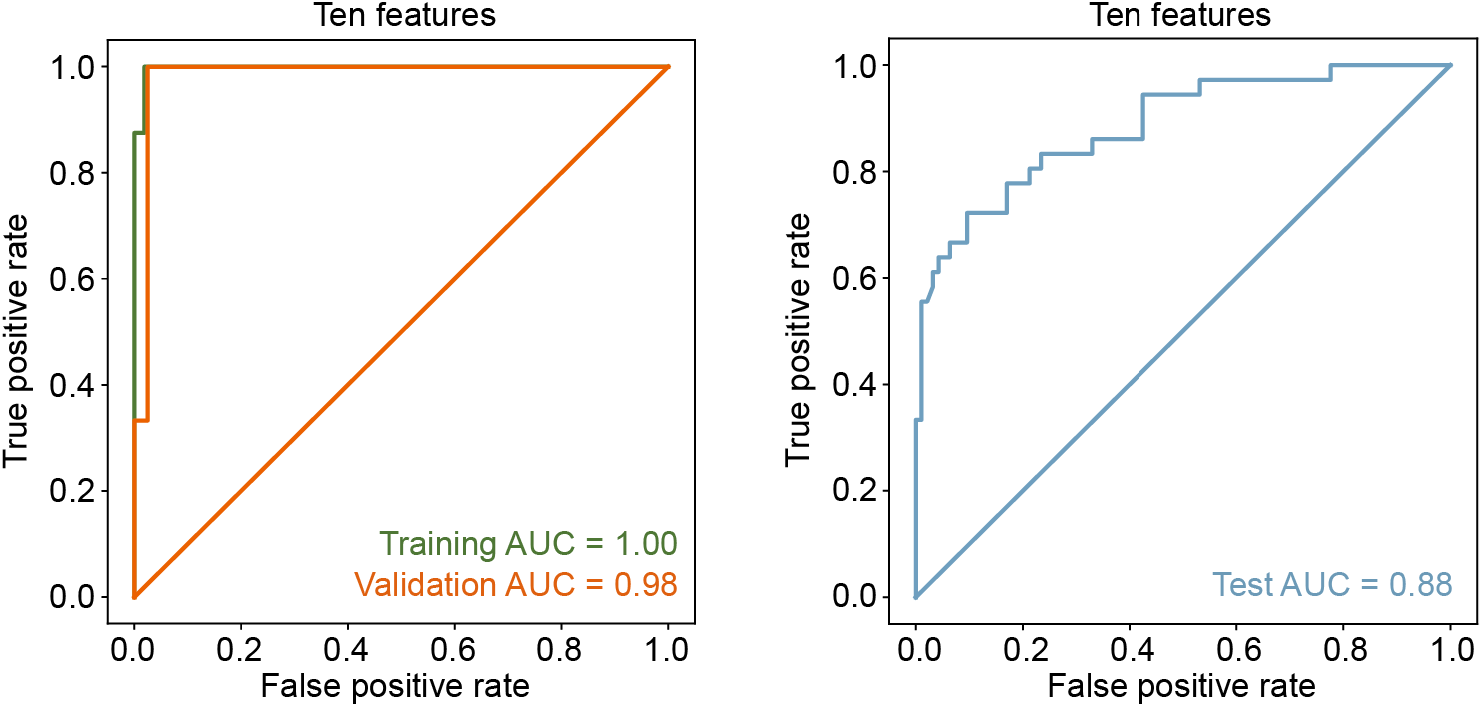
Performance of the ten-feature classifier without feature Mg. ROC plots the performance of support vector machine for severe cases prediction using same ten clinical features mentioned in main text without Mg. The AUC was decreased by 0.01 compared with eleven features.

**Table S1.**
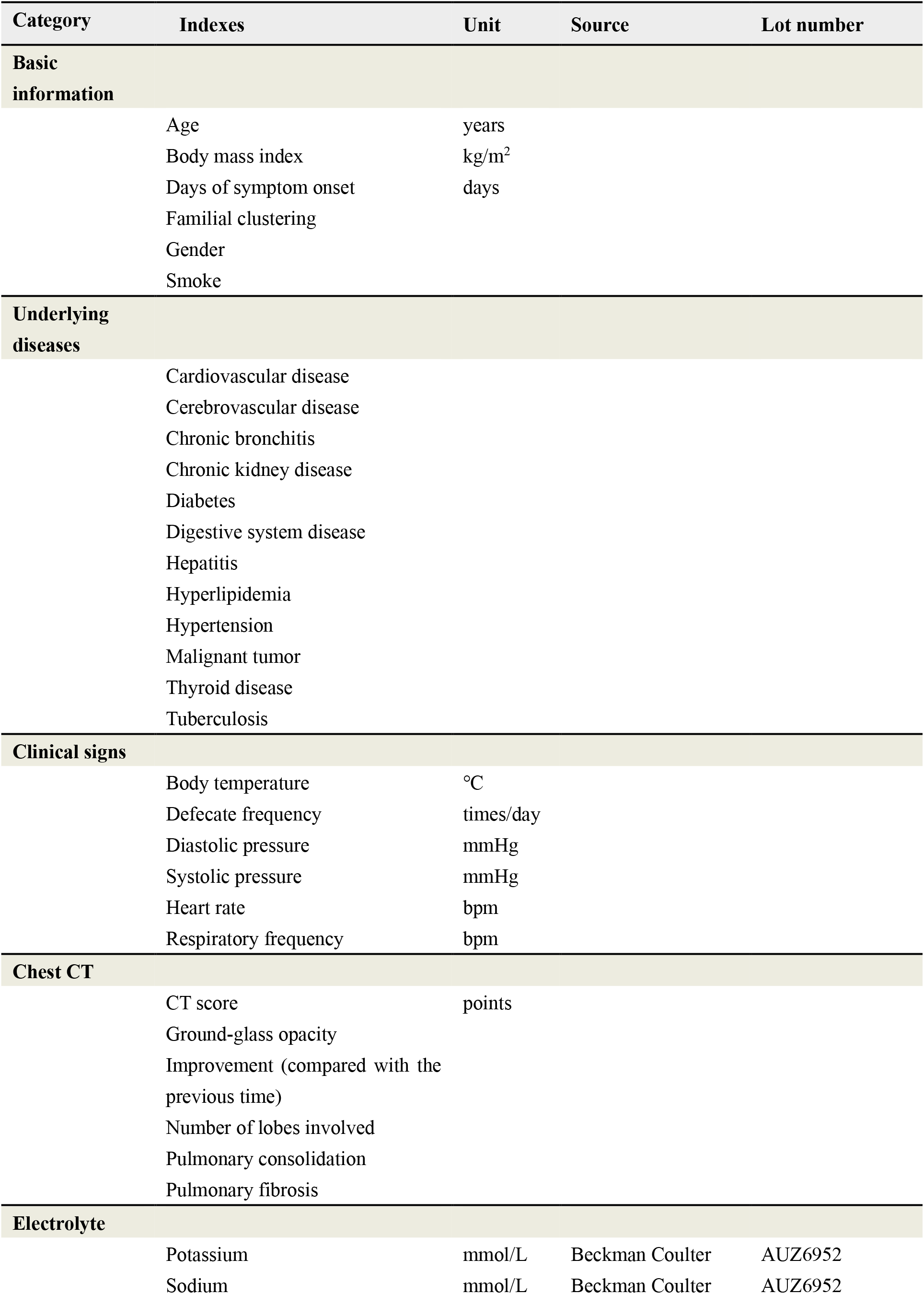
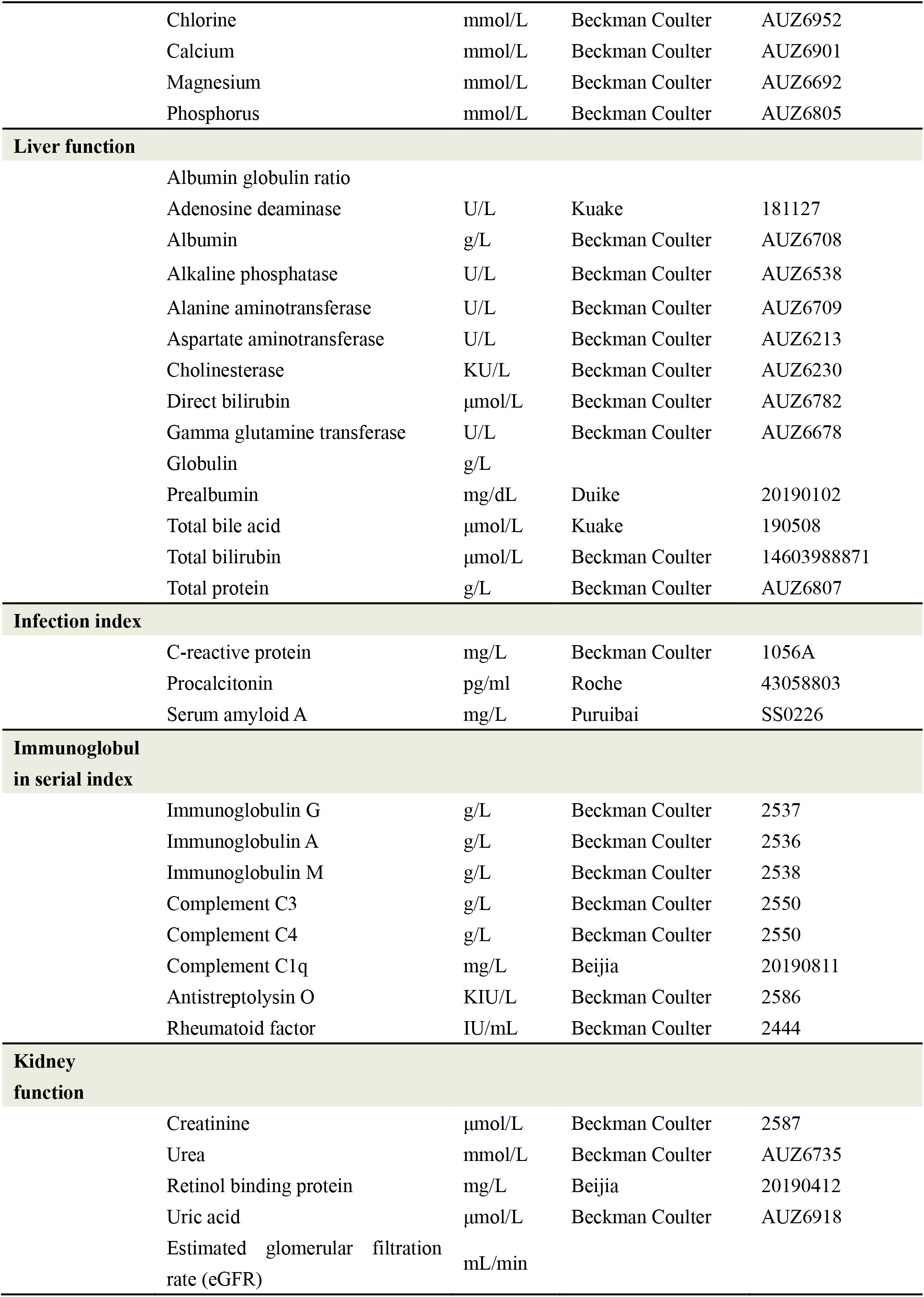
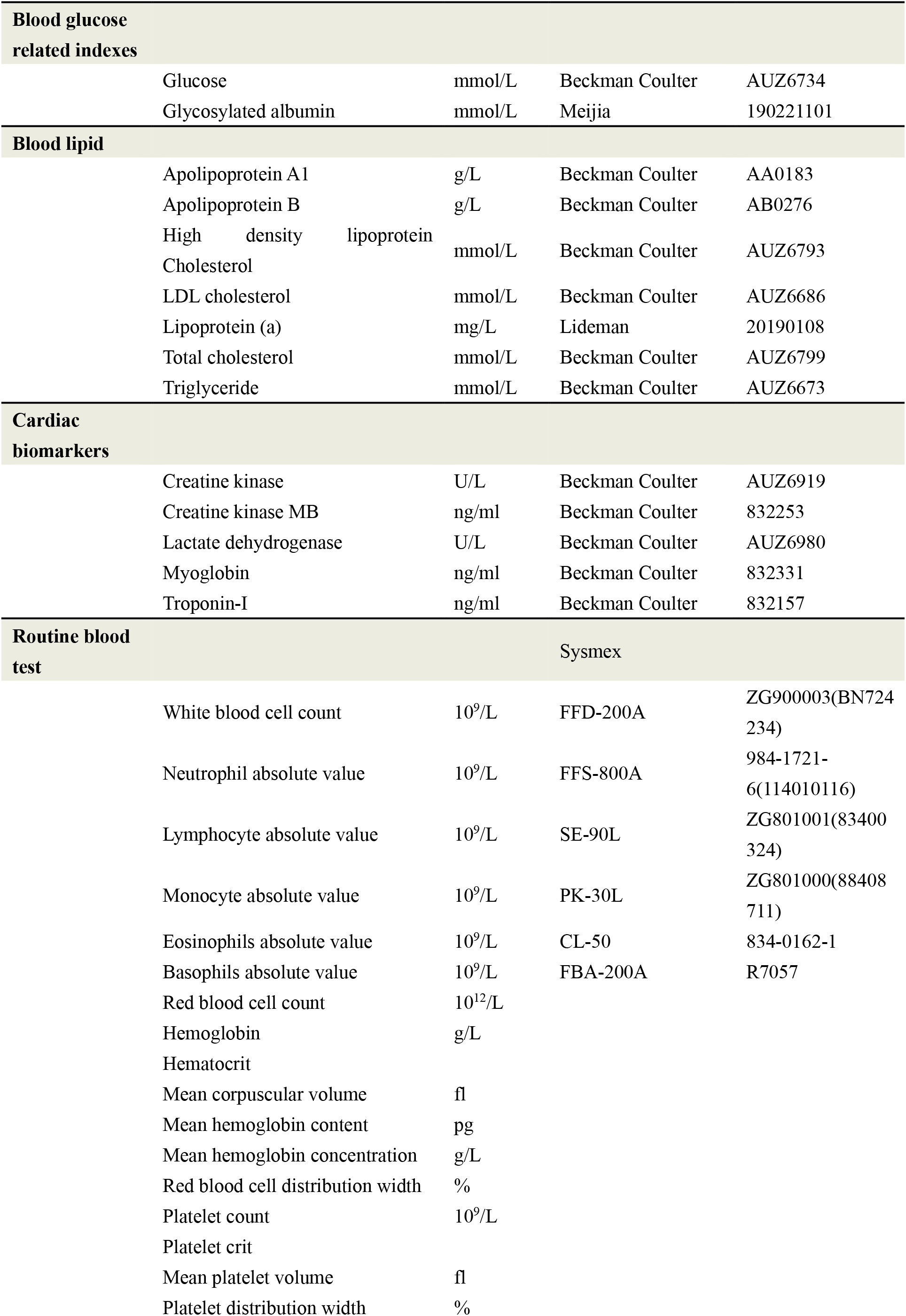
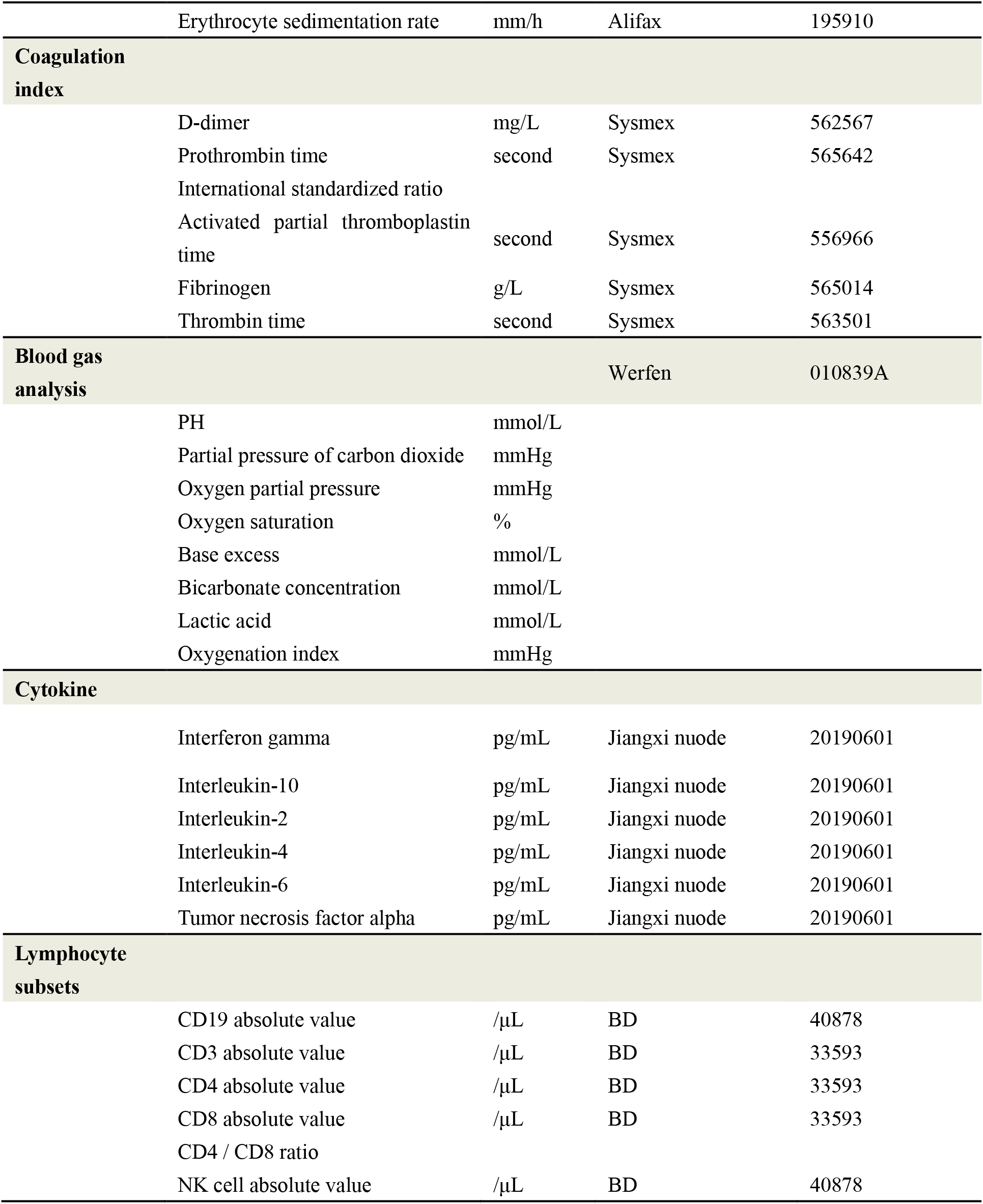
17 categories of indexes.

